# Effect of Screen time on Glycaemic control of Type 2 Diabetes patients during COVID-19 Outbreak: A Survey based Study

**DOI:** 10.1101/2020.09.09.20188961

**Authors:** Sayak Roy, Kingshuk Bhattacharjee

**Affiliations:** Medica Superspeciality Hospital, Kolkata, India; Medwes Pvt LTD, Kolkata, Wb, India

**Keywords:** Coronavirus disease 2019, glycosylated hemoglobin, diabetes mellitus type 2, social media

## Abstract

**Introduction:** Decreased workout during coronavirus disease 2019 (COVID-19) is a serious issue for the patients with type 2 diabetes (T2DM), since their glycaemic control is very much related to that. COVID-19 has posed a severe health issue that is playing havoc on the aged patients with existing comorbidities. Studies have shown mixed reports of social media on T2DM, with some showing positive results due to increased use of apps and adherence to lifestyle, while others have shown adiposity and glycaemic control related to hours spent on-screen time in children. Data on adult T2DM patients’ screen time activity and prevailing glycosylated haemoglobin (HbA1c), fasting blood sugar (FBS) and, post-prandial blood sugar (PPBS) is sparse.

**Aim:** To study the effect of screen-time spent on social media per day on glycaemic parameters of T2DM patients.

**Materials and methods:** Data was collected for T2DM patients giving informed written consent and meeting a set of pre-specified inclusion criteria. Through two rounds of surveys done from May 15 to June 26, the authors collected the answers to a set of questionnaires from a total of 344 patients sent via email. Due to the non-availability of data from a few patients, a total of 229 patients’ data were finally analyzed. SPSS software version V21 ® was used to perform Binary logistic regression for calculating the odds ratio (OR) of the categorical variables. The outcomes, looked for in the analysis, were poor control of glycaemic parameters like HbA1c (defined by >7%), FBS (defined by >150 mg/dL) and, PPBS (defined by >200 mg/dL) and the exposure variables were Screen time spent by the person per day for all the three glycaemic parameters and, doctor’s visit and, daily exercise for HbA1c outcome.

**Results:** A total of 173 patients had a screen time (henceforth, it means time spent on social media) of less than 2 hours/day in the study sample. Among the 173 patients, 73 (42.2%) had achieved HbA1c less than 7%, whereas the remaining 100 (57.8%) had HbA1c more than 7%. On the other hand, 56 patients had a screen time of more than 2 hours, of which 44 (72.73%) had HbA1c more than 7%. Among the 173 patients, only 89 (51.44%) had an FBS value of more than 150 mg/dL as compared to 46 (82.12%) with a screen time of more than 2 hours. Out of these 173 patients, only 43 (24.86%) had a PPBS value of more than 200 mg/dL as compared to 41 (73.21%) with a screen time of more than 2 hours. It was found that the odds of having a poor glycaemic control as per HbA1c, FBS and PPBS is 2.67 times higher (95%CI: 1.91-6.95), 4.34 times higher (95%CI: 1.52-4.76) and, 8.26 times higher (95%CI: 4.26-11.83) in the cohort with a screen time of more than 2 hours as compared to the cohort with a screen time of less than 2 hours, respectively.

**Conclusion:** There seems to be an increased risk of uncontrolled glycaemic indices with increased screen time and, decreased work out. This is a small study and the findings need to be corroborated with larger sample size.

## Introduction

Screen-time has been linked to adiposity and insulin resistance [1]. Evidence suggests CVD risk factors are present to a significantly higher degree in the subset of overweight/obese individuals that are also insulin resistant [2]. A multivariate analysis, after adjusting for smoking, age, exercise levels per day and other lifestyle factors in a prospective cohort study of 6 years, showed that there was 23% (95% confidence interval [CI], 17%-30%) increase in obesity with each 2-h/day increment in TV watching and a 14% increase in risk of developing diabetes [**3**]. There has been elevated all-cause and cardiovascular disease mortality risk, even after physical activity adjustment, with increased sitting time/sedentary behaviours [**4**]. The outbreak of COVID-19 has led to reduced physical activity because of higher chances of getting infection on venturing outside home [**5**]. It is presumed that prolonged staying at home due to this COVID-19 will increase sedentary habits like increased time spent on mobile phones and watching TV [**6**]. The WHO has set a target time to be spent for exercise in adults and the elderly, which is estimated to be 150 min/week of moderate physical activity [**7**].

Taking this idea into consideration, a cross-sectional survey-based analysis was conducted to see the effect of screen time spent by a T2DM patient during this COVID-19 pandemic on the glycemic parameters.

## Materials and methods

This was a cross sectional study in which data were collected through two rounds of surveys (with answer options) sent to regular outdoor T2DM patients visiting the authors’ clinics from May 15 to June 26, 2020, via patients’ pre-registered email IDs. The local ethical body to which the authors are attached (*Remedy Clinic Study Group*) approved the study.

There is no ready set of questionnaire available on association between social media and glycaemia during this lockdown period and hence, the authors used a new set of questionnaires in English language, the idea of which were taken from the Diabetes Self-Management Questionnaire (DSMQ) [**8**], the Maastricht Study [**9**] and, also from a Saudi Arabian study [**10**].The authors validated their questionnaires by three independent expert physicians.

Patients’ last visits’ electronic health record was available in the author’s clinic database to look for the patients who were previously compliant with follow-up and medicines and those patients who did mention their previous social media time spent use, as a routine questionnaire in clinic, less than 1 hour were included. The patients matching baseline inclusion criteria were contacted telephonically before the emails were sent. Since many patients were not able to come to regular check-ups during the COVID-19 outbreak, but many did their routine tests, questionnaires were shared to collect the data.

The questions were provided with easy and flexible answer options (using categorical variables) for the ease of the patients who might not be able to write down the exact values. After contacting the selected group of patients telephonically, emails were sent for written informed consent form along with the survey questionnaires (figure 1). The questionnaires were distributed via email and patients were asked to revert within 10 days.

**Figure 1.**
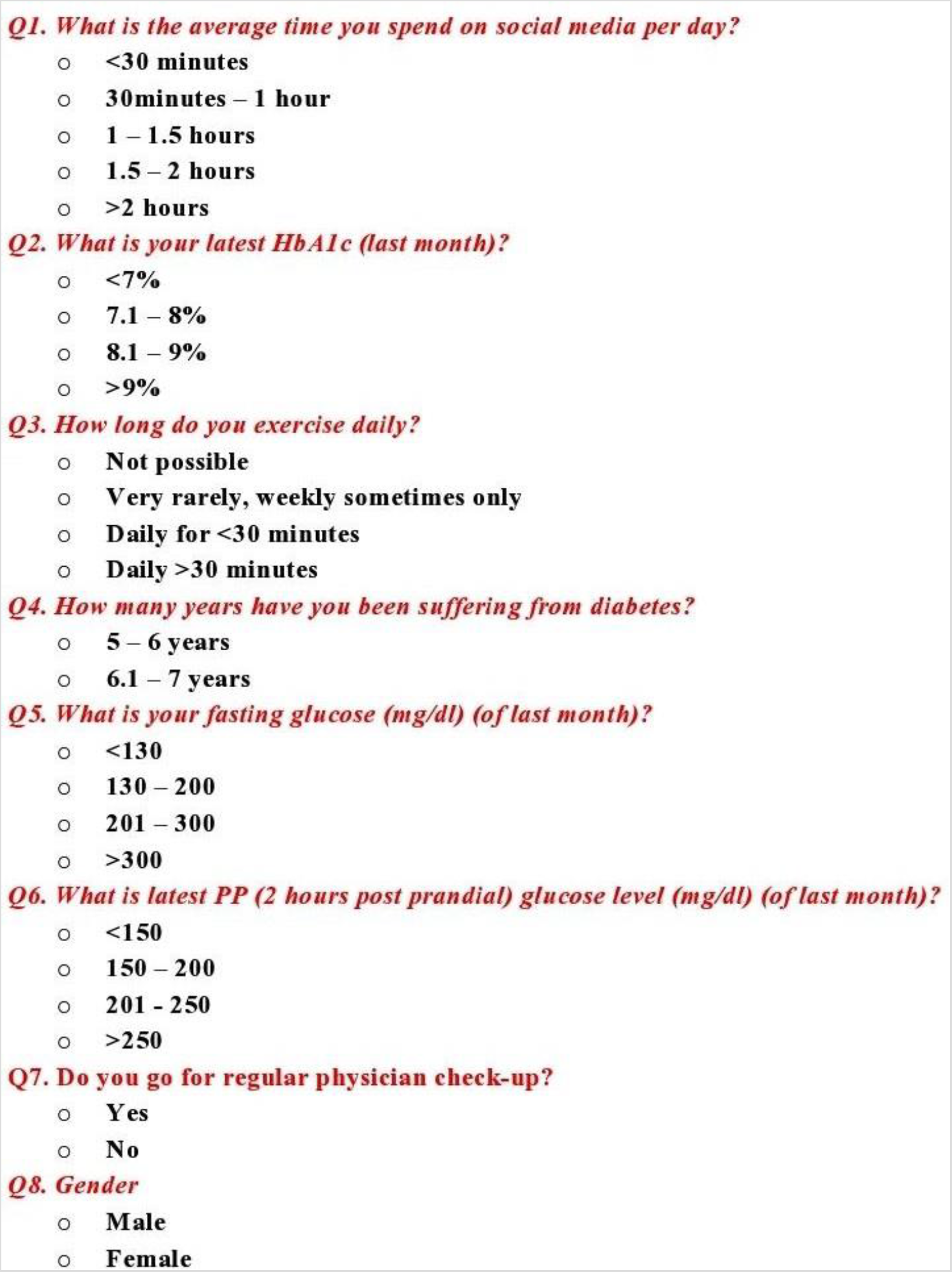
Survey questionnaires with answers

Inclusion criteria: a) T2DM patients; b) BMI 20 – 28 Kg/m^2^, c) age 19 – 60 years; d) estimated glomerular filtration rate (eGFR) as per CKD-EPI of >60 ml/min/1.73m^2^; d) duration of diabetes 5 – 7 years and, f) No injectable in existing treatment regimen. Exclusion criteria: a) if there was any history of hospitalization in last three months and, b) if they did not give informed consent to share data.

A total of 344 (first round of the survey – 158 participants; the second round of the survey – 186 participants) patients participated. The first round of the survey was carried out from May 15 to May 29, and the second-round survey was carried out from June 15 to June 26, 2020. In the first round of the survey, 50 patients did not mention one of their glycemic parameters like HbA1c, FBS, or PPBS, and in the second round, 65 patients did not report any one of the above parameters. Hence their data were excluded from the final analysis. Finally, data of 108 patients from round one of the survey and 121 patients’ data from the second round of the survey were available for analysis. The questions used in the survey with their answer options are given in the appendix.

## Sample size calculation

Due to paucity of published data related to the present study on the effect of screen time on glycaemic status in the intended study population from Eastern India, the sample size could not be computed by conventional sample size formula. Hence the sample size is based on the Thumb’s rules. When no baseline information for computation of sample size is available, Thumb rule states a minimum of 120 subjects for a cross-sectional survey. This sample size is bare minimum to carry out statistical data analysis after completion of the study [**11**].

## Statistical analysis

SPSS software version V21 ® was used to perform Binary logistic regression for calculating the Odds ratio (OR) of the categorical variables. The outcomes, looked for in the study, were poor control of glycemic parameters like HbA1c (defined by >7%), FBS (defined by >150 mg/dL) and, PPBS (defined by >200 mg/dL) and the exposure variables were screen time spent by the person per day for all the three glycemic parameters, doctor’s visit and daily exercise for HbA1coutcome. Descriptive analysis was performed, and the data were expressed as numbers and percentages. Comparison between the proportion of the study parameters before and during the lockdown has been carried out by extended McNemar test.

## Results

The baseline characteristics of the total analysed cohort is summarised in table 1.

**Table 1.** Characteristics of the study population during lockdown

| Parameters | Number (%) | Total |
| --- | --- | --- |
| **Gender** |  | 229 |
| Male | 100 (43.67) |  |
| Female | 129 (56.33) |  |
| **Mean Age with standard deviation** |  | 229 |
| Male (years) (n=100) | 42.2 ±2.516 (±5.96%) |  |
| Female (years) (n=129) | 44.2093 ±2.183 (±4.94%) |  |
| **Duration of Diabetes** |  | 229 |
| 5 – 6 years | 67 (29.26) |  |
| 6.1 – 7 years | 162 (70.74) |  |
| **Time spent on social media** |  | 229 |
| Less than 30 minutes | 34 (14.85) |  |
| 30 minutes- 1 hour | 43 (18.78) |  |
| 1 hour – 1.5 hours | 54 (23.8) |  |
| 1.5 hours- 2 hours | 42 (18.34) |  |
| More than 2 hours | 56 (24.45) |  |
| **Daily aerobic exercise time** |  | 229 |
| Not possible | 23 (10.04) |  |
| Very rarely | 82 (35.81) |  |
| Less than 30 minutes | 49 (21.4) |  |
| More than 30 minutes | 75 (32.75) |  |
| **Physician check-up (as per ADA guideline) [12]** |  | 229 |
| Regular (defined as every 3 months if uncontrolled and, every 6 months if controlled) | 191 (83.41) |  |
| Not Regular | 38 (16.59) |  |
| **Latest HbA1c (glycated haemoglobin) (within last month)** |  | 229 |
| Less than 7% | 85 (37.12) |  |
| 7.1% - 8% | 67 (29.25) |  |
| 8.1% - 9% | 44 (19.21) |  |
| Greater than 9% | 33 (14.41) |  |
| **Fasting Blood Sugar (within last month)** |  | 229 |
| Less than 130 mg/dL | 79 (34.50) |  |
| 130-200 mg/dL | 74 (32.31) |  |
| 201-300 mg/dL | 35 (15.28) |  |
| More than 300 mg/dL | 41 (17.9) |  |
| **Post-prandial Blood Sugar (within last month)** |  | 229 |
| Less than 150 mg/dL | 59 (25.76) |  |
| 150-200 mg/dL | 86 (37.55) |  |
| 201-250 mg/dL | 44 (19.21) |  |
| More than 250 mg/dL | 40 (17.46) |  |

The changes of the patients between pre-lockdown period and lockdown period is calculated by extended McNemar test which shows the glycaemic indices during lockdown period to be significantly higher than previous state and are depicted in table 2.

**Table 2.**
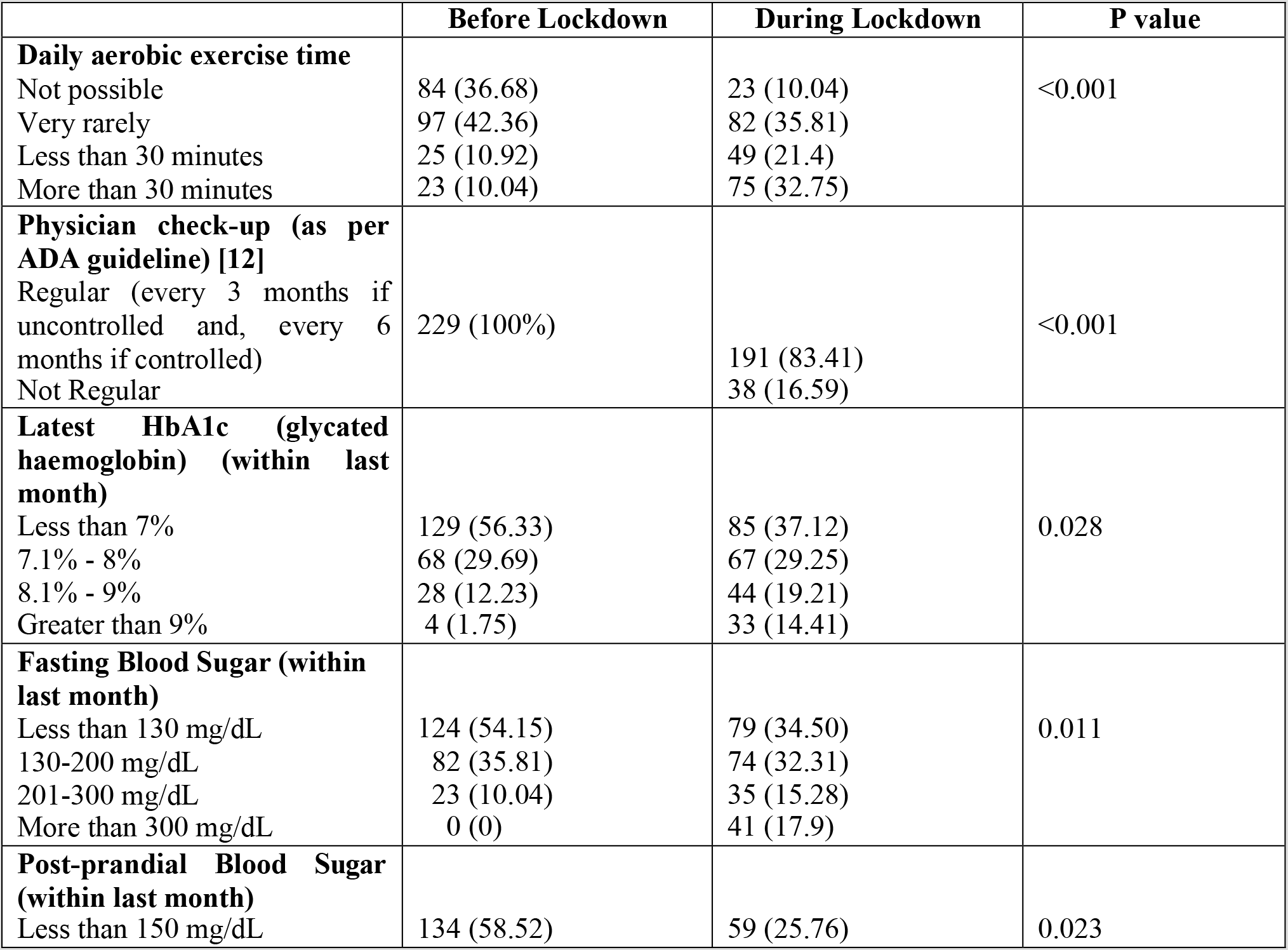
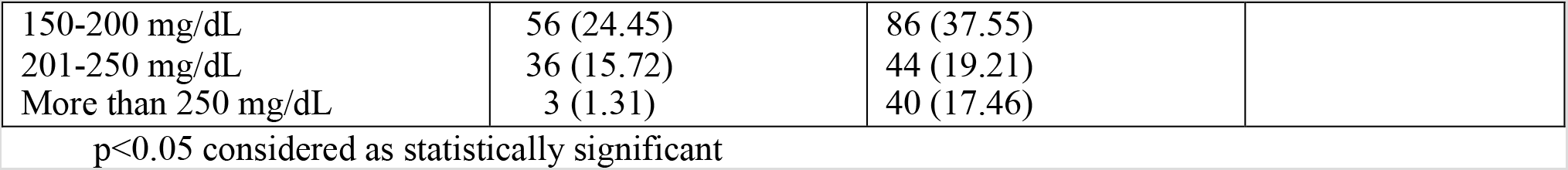
Changes in the study parameters before and after lockdown

|  | Before Lockdown | During Lockdown | P value |
| --- | --- | --- | --- |
| **Daily aerobic exercise time** |  |  |  |
| Not possible | 84 (36.68) | 23 (10.04) | <0.001 |
| Very rarely | 97 (42.36) | 82 (35.81) |  |
| Less than 30 minutes | 25 (10.92) | 49 (21.4) |  |
| More than 30 minutes | 23 (10.04) | 75 (32.75) |  |
| **Physician check-up (as per ADA guideline) [12]** |  |  |  |
| Regular (every 3 months if uncontrolled and, every 6 months if controlled) | 229 (100%) | 191 (83.41) | <0.001 |
| Not Regular |  | 38 (16.59) |  |
| **Latest HbA1c (glycated haemoglobin) (within last month)** |  |  |  |
| Less than 7% | 129 (56.33) | 85 (37.12) | 0.028 |
| 7.1% - 8% | 68 (29.69) | 67 (29.25) |  |
| 8.1% - 9% | 28 (12.23) | 44 (19.21) |  |
| Greater than 9% | 4 (1.75) | 33 (14.41) |  |
| **Fasting Blood Sugar (within last month)** |  |  |  |
| Less than 130 mg/dL | 124 (54.15) | 79 (34.50) | 0.011 |
| 130-200 mg/dL | 82 (35.81) | 74 (32.31) |  |
| 201-300 mg/dL | 23 (10.04) | 35 (15.28) |  |
| More than 300 mg/dL | 0 (0) | 41 (17.9) |  |
| **Post-prandial Blood Sugar (within last month)** |  |  |  |
| Less than 150 mg/dL | 134 (58.52) | 59 (25.76) | 0.023 |
| 150-200 mg/dL | 56 (24.45) | 86 (37.55) |  |
| 201-250 mg/dL | 36 (15.72) | 44 (19.21) |  |
| More than 250 mg/dL | 3 (1.31) | 40 (17.46) |  |

$p < 0.05$ considered as statistically significant

A total of 173 patients had a screen time of less than 2 hours/day in our study sample. Among the 173 patients, 73 (42.2%) had achieved HbA1c less than 7%, whereas the remaining 57.8% had HbA1c more than 7%. On the other hand, 56 patients had a screen time of more than 2 hours, out of which 44 (72.73%) had HbA1c more than 7%. Among the 173 patients, only 89 (51.44%) had an FBS value of more than 150 mg/dL as compared to 46 (82.12%) with a screen time of more than 2 hours. Out of these 173 patients, only 43 (24.86%) had a PPBS value of more than 200 mg/dL as compared to 41 (73.21%) with a screen time of more than 2 hours. By conducting binary logistic regression, we found that the odds of having a poor glycaemic control as per HbA1c, FBS and PPBS is 2.67 times higher (95%CI: 1.91-6.95), 4.34 times higher (95%CI: 1.52-4.76) and, 8.26 times higher (95%CI: 4.26–11.83) in the cohort with a screen time of more than 2 hours as compared to the cohort with a screen time of less than 2 hours, respectively (table 3).

**Table 3.** Odds ratio of poor glycaemic control parameters (outcome parameters) – HbA1c, FBS and PPBS according to screen time (exposure parameter)

| *HbA1c* |  |  |  |  |
| --- | --- | --- | --- | --- |
| Screen time (hour/day) | < 7 % | >7% | Odds Ratio (OR) | 95% Confidence Interval |
| < 2 hours; n (%) | 73 (42.2%) | 100 (57.8%) | 1.00 | Reference |
| >2 hours; n (%) | 12 (27.27%) | 44 (72.73%) | 2.67 | 1.91 – 6.95 |
| *FBS* |  |  |  |  |
| Screen time (hour/day) | <150 mg/dL | >150 mg/dL | Odds Ratio (OR) | 95% Confidence Interval |
| < 2 hours; n (%) | 84 (48.56%) | 89 (51.44%) | 1.00 | Reference |
| >2 hours; n (%) | 10 (17.88%) | 46 (82.12%) | 4.34 | 6.63-14.16 |
| *PPBS* |  |  |  |  |
| Screen time (hour/day) | <200 mg/dL | >200 mg/dL | Odds Ratio | 95% Confidence Interval |
| < 2 hours; n (%) | 130 (75.14) | 43 (24.86) | 1.00 | Reference |
| >2 hours; n (%) | 15 (26.79) | 41 (73.21) | 8.26 | 4.26 – 11.83 |

*OR* > 1, Exposure associated with higher odds of outcome; *HbA1c*, glycated haemoglobin; *FBS*, fasting blood sugar; *PPBS*, post-prandial blood sugar

The OR calculated for HbA1c (outcome variable) taking exposure variables of daily exercise time >30 minutes/day as standard cut-off and regular doctors’ visits, were 3.91 (95%CI: 1.22-2.76) and 2.73 (95%CI: 1.12-2.98), respectively. This means that the cohort having spent less than 30 minutes/day exercising had 3.91 times higher odds of having poor HbA1c control, and those not going for regular check-up also had a 2.73 higher odds of not achieving target HbA1c of < 7% (table 4).

**Table 4.** Odds ratio of poor HbA1c control (outcome parameter) according to regular exercise (>30 minutes/day) and regular physician check-up (exposure parameter)

| *Exercise* |  |  |  |  |
| --- | --- | --- | --- | --- |
| Exercise (minutes/day) | *HbA1c* |  | Odds Ratio | 95% Confidence Interval |
|  | < 7 % | >7% |  |  |
| < 30 minutes; n (%) | 41 (26.62) | 113 (73.38) | 1.00 | Reference |
| > 30 minutes; n (%) | 44 (58.67) | 31 (41.33) | 3.91 | 1.22 – 2.76 |
| *Regular physician check up* |  |  |  |  |
| Regular check-up | *HbA1c* |  | Odds Ratio | 95% Confidence Interval |

|  | < 7 % | >7% |  |  |
| --- | --- | --- | --- | --- |
| **Yes; n (%)** | 79 | 154 | 1.00 | *Reference* |
| **No; n (%)** | 6 | 32 | 2.73 | 1.12-2.98 |

*OR>1, Exposure associated with higher odds of outcome; HbA1c, glycated haemoglobin*

## Discussion

Covid-19 has considerably changed our lifestyle and has reduced our physical activity [**5**], leading to increased sedentary habits [**6**]. This study tried to shed some light on an entirely new dimension of COVID-19 induced reduced physical activity, increased screen time, and their effect on HbA1c and other glycemic parameters. We deliberately took a duration of diabetes of 5 – 7 years since that will help us get a population where there will be a chance of optimizing oral anti-diabetic drugs (OADs), and hence the inclusion criteria of no injectable can be justified. The age criteria of 19 – 60 years relate to an active, healthy cohort who can carry out exercise or lifestyle changes as per physician advice. It has been found that only 54.6% of Indian T2DM patients comply with dietary advice, and only 37.2% follow the advice of exercise [**13**]. A meta-analysis of controlled clinical trials have shown a significant reduction of HbA1c in the arm doing exercise V/s. the control group (7.65% V/s. 8.31%; weighted mean difference, –0.66%; P<.001) [14]. Another meta-analysis found no significant difference between aerobic exercise and resistance exercise after sensitivity analysis (p = 0.14) [**15**]. Interleukin 6 (IL-6) and C-Reactive Protein (CRP) have been used as surrogate systemic inflammatory markers associated with T2DM [**16**]. A study with 60 obese women undergoing lifestyle modifications by physical activity and weight loss, showed a marked reduction in inflammatory markers like IL-6 (p = 0.009), interleukin 18 (IL-18) (p = 0.02) and, CRP (p = 0.008) [**17**].

In this era of mobile devices, text messages and telephonic communications play a significant role, and studies have demonstrated a similar efficacy of short message service (SMS) by cellular phone and telephonic follow-up by a nurse in reducing HbA1c [**18**]. A prospective, parallel-group, randomized controlled trial conducted in Asian Indian men with impaired glucose tolerance showed a significant decrease in developing overt T2DM in the intervention group, which received lifestyle advice at regular intervals via SMS (n = 271) than the control arm (n = 266), p-value 0.015 [**19**]. Similarly, a web-based survey done on both type 1 (n = 549) and type 2 diabetes (n = 210) found a significantly high self-care behavior score with diabetes app users after adjustment of confounding factors, for type 1 the increase was 1.08 (95%CI: 0.46-1.7) units and for T2DM it increased by 1.18 (95%CI: 0.26-2.09) units [**20**]. Not only positive but also negative impacts are seen in many systematic reviews that are done, and we often get mixed responses [**21**].

This study found an increase in HbA1c, FBS and, PPBS with increased time spent on social media, reduced physical activity, and irregular physician follow-up.

## Study limitations

There are many limitations in this study:

1. As a survey-based study, we had to depend solely on the patient’s data and could not verify it.
2. BMI could not be adjusted since it was not included in the answer sheet and, the electronic health record had the last visit BMI which might have changed during the survey time due to a sedentary lifestyle during the lockdown period.
3. We had to use a range for all the questions (categorical variable) since patients find it easy to answer, hence we could not get the exact value of all parameters and thus could not do a multiple linear regression analysis to look for adjustments or other variables that could have affected the outcome.

## Conclusion

Since the outbreak of this pandemic, our life changed to a considerable extent. We need to tackle this problem with new ways as per suggestions by the international bodies, and T2DM patients should carry on their daily activity at home or in nearby areas taking proper recommended precautions. Rare papers are there on screen time spent and glycaemic outcomes in T2DM patients during this lockdown period. There seems to be an increased risk of uncontrolled glycaemia if the patient spends more time on social media with reduced physical activity, without any productive events, and these sort of sedentary changes are enhanced in these types of outbreaks. Social media can be used in many useful ways to communicate with the patients and provide awareness, but the non-judicious use might become a problem.

## Data Availability

Available on request with the authors.

## Acknowledgement

None

## References

1. Nightingale CM, Rudnicka AR, Donin AS, et al. Screen time is associated with adiposity and insulin resistance in children. Arch Dis Child. 2017; 102(7): 612-616. doi: 10.1136/archdischild-2016-312016

2. Reaven GM. Insulin resistance: the link between obesity and cardiovascular disease. Med Clin North Am. 2011; 95(5): 875-892. doi: 10.1016/j.mcna.2011.06.002

3. Hu FB, Li TY, Colditz GA, Willett WC, Manson JE. Television watching and other sedentary behaviors in relation to risk of obesity and type 2 diabetes mellitus in women. jama. 2003; 289(14): 1785-1791. doi: 10.1001/jama.289.14.1785

4. Shiyovich A, Shlyakhover V, Katz A. Harefuah. 2013; 152(1): 43-57 [Sitting and Cardiovascular Morbidity and Mortality]. Available at: https://pubmed.ncbi.nlm.nih.gov/23461028/?from_term=screen+time+spent+and+type+2+diabetes&from_filter=pubt.review&from_pos=2 [Accessed 26 May 2020]

5. Kim, S.-W. and Su, K.-P. (2020). Using psychoneuroimmunity against COVID-19. Brain, Behavior, and Immunity. doi: 10.1016/j.bbi.2020.03.025 [Epub ahead of print]

6. Chen, P., Mao, L., Nassis, G.P., et al. (2020). Coronavirus disease (COVID-19): The need to maintain regular physical activity while taking precautions. Journal of Sport and Health Science, 9(2), pp.103–104. doi: 10.1016/j.jshs.2020.02.001

7. AmriHammami, BasmaHarrabi, Magni Mohr & Peter Krustrup (2020): Physicalactivity and coronavirus disease 2019 (COVID-19): specific recommendations for home-basedphysical training, Managing Sport and Leisure, DOI: 10.1080/23750472.2020.1757494

8. Schmitt, A., Gahr, A., Hermanns, N., Kulzer, B., Huber, J. and Haak, T. (2013). The Diabetes Self-Management Questionnaire (DSMQ): development and evaluation of an instrument to assess diabetes self-care activities associated with glycaemic control. Health and Quality of Life Outcomes, [online] 11(1), p.138. Available at: https://www.ncbi.nlm.nih.gov/pmc/articles/PMC3751743/ [Accessed 25 Nov. 2019]. doi: 10.1186/1477-7525-11-138

9. Brinkhues, S., Dukers-Muijrers, N.H.T.M., Hoebe, C.J.P.A., van der Kallen, C.J.H., Koster, A., Henry, R.M.A., Stehouwer, C.D.A., Savelkoul, P.H.M., Schaper, N.C. and Schram, M.T. (2018). Social Network Characteristics Are Associated With Type 2 Diabetes Complications: The Maastricht Study. Diabetes Care, 41(8), pp.1654–1662. https://doi.org/10.2337/dc17-2144

10. Alzahrani, A. and Alanzi, T. (2019). Social Media Use By People With Diabetes In Saudi Arabia: A Survey About Purposes, Benefits And Risks. Diabetes, Metabolic Syndrome and Obesity: Targets and Therapy, Volume 12, pp.2363–2372. https://doi.org/10.2147/DMSO.S208141

11. AbhayaIndrayan (2013). Medical biostatistics. Boca Raton, Confidence Intervals, Principles of Tests of Significance, and sample size. Fla. London: Chapman & Hall/CrcBiostatistics Series; 2013: 423-443.

12. 6. Glycemic Targets: Standards of Medical Care in Diabetes—2020. (2019). Diabetes Care, 43(Supplement 1), pp.S66–S76. https://doi.org/10.2337/dc20-S006

13. Mohan V, Shah SN, Joshi SR, et al., On Behalf of the DiabCare India 2011 Study Group. Current status of management, control, complications and psychosocial aspects of patients with diabetes in India: Results from the DiabCare India 2011 Study. Indian J EndocrMetab 2014; 18: 370-8, DOI: 10.4103/2230-8210.129715

14. Boulé NG, Haddad E, Kenny GP, Wells GA, Sigal RJ. Effects of exercise on glycemic control and body mass in type 2 diabetes mellitus: a meta-analysis of controlled clinical trials. JAMA. 2001; 286(10): 1218-1227. doi: 10.1001/jama.286.10.1218

15. Yang Z, Scott CA, Mao C, Tang J, Farmer AJ. Resistance exercise versus aerobic exercise for type 2 diabetes: a systematic review and meta-analysis. Sports Med. 2014; 44(4): 487-499. doi: 10.1007/s40279-013-0128-8

16. Festa A, D’Agostino R Jr, Howard G, Mykkänen L, Tracy RP, Haffner SM. Chronic subclinical inflammation as part of the insulin resistance syndrome: the Insulin Resistance Atherosclerosis Study (IRAS). Circulation. 2000; 102(1): 42-47. doi: 10.1161/01.cir.102.1.42

17. Esposito K, Pontillo A, Di Palo C, et al. Effect of weight loss and lifestyle changes on vascular inflammatory markers in obese women: a randomized trial. JAMA. 2003; 289(14): 1799-1804. doi: 10.1001/jama.289.14.1799

18. Zolfaghari, M., Mousavifar, S. A., & Haghani, H. (2012). Mobile phone text messaging and Telephone follow-up in type 2 diabetic patients for 3 months: a comparative study. Journal of diabetes and metabolic disorders, 11(1), 7. https://doi.org/10.1186/2251-6581-11-7 (Retraction published J Diabetes MetabDisord. 2013 Mar 07;: 11(1): 21)

19. Ramachandran, A., Snehalatha, C., Ram, J., Selvam, S., Simon, M., Nanditha, A., Shetty, A.S., Godsland, I.F., Chaturvedi, N., Majeed, A., Oliver, N., Toumazou, C., Alberti, K.G. and Johnston, D.G. (2013). Effectiveness of mobile phone messaging in prevention of type 2 diabetes by lifestyle modification in men in India: a prospective, parallel-group, randomised controlled trial. The Lancet Diabetes & Endocrinology, 1(3), pp.191–198. https://doi.org/10.1016/S2213-8587(13)70067-6

20. Kebede MM, Pischke CR. Popular Diabetes Apps and the Impact of Diabetes App Use on Self-Care Behaviour: A Survey Among the Digital Community of Persons With Diabetes on Social Media [published correction appears in Front Endocrinol (Lausanne). 2019 Apr 05;: 10: 220]. Front Endocrinol (Lausanne). 2019; 10: 135. Published 2019 Mar 1. doi: 10.3389/fendo.2019.00135

21. Giustini D, Ali SM, Fraser M, KamelBoulos MN. Effective uses of social media in public health and medicine: a systematic review of systematic reviews. Online J Public Health Inform. 2018; 10(2): e215. Published 2018 Sep 21. doi: 10.5210/ojphi.v10i2.8270

